# Assessing Attention Process Training Efficacy in Improving Brain Fog Symptoms in Individuals with Long Covid: A Study Protocol for a Randomized Controlled Trial

**DOI:** 10.64898/2026.03.10.26348047

**Authors:** Kathryn Magee, Elliot Roth, Leora R. Cherney, Shira Cohen-Zimerman

## Abstract

**Background:** Long Covid, also referred to as post-acute sequelae of SARS-CoV-2 infection (PASC), is characterized by symptoms that persist or emerge weeks to months following acute COVID-19 illness. Cognitive impairments affecting attention, memory, and executive functioning—commonly described as “brain fog”—are frequently reported. Currently, limited evidence-based cognitive rehabilitation interventions specifically target these impairments.

**Objectives:** This pilot randomized controlled trial aims to evaluate the feasibility and acceptability, and to develop preliminary data regarding efficacy of Attention Process Training–3 (APT-3), a computerized attention training program, for individuals with Long Covid–related brain fog.

**Methods:** This study uses a three-arm randomized controlled design. Participants are randomized to (1) Immediate Attention Training, (2) Delayed Attention Training, or (3) Music Activity. Participants complete comprehensive cognitive assessments at baseline, post-intervention, and one-month follow-up. The Immediate Attention Training group completes a 4-week APT-3 intervention, while the Music Activity group engages in a 4-week music-based listening activity. The Delayed Attention Training group don’t receive any intervention for 4 weeks. Following completion of the final assessment, participants in the Music Activity and Delayed Attention Training groups are offered the APT-3 intervention. Feasibility and acceptability outcomes include recruitment, retention, and adherence numbers, and participant satisfaction. Preliminary data regarding efficacy will be determined using objective cognitive tests and subjective self-report measures.

**Conclusions:** This pilot trial will inform the feasibility and acceptability of APT-3 and generate preliminary efficacy data to guide the design of a future fully powered randomized controlled trial targeting brain fog associated with Long Covid.

## Background

The diagnosis of Long Covid reflects the constellation of post-acute and long-term health effects caused by SARS-CoV-2 infection; it is a complex, multisystem disorder that can affect nearly every organ system and can be severely disabling (1). It is well established that Long Covid remains a significant health challenge (2). A substantial proportion of people with Long Covid report cognitive impairments, particularly in attention, memory and executive functions (3–8).

These impairments are commonly referred to as “*brain fog”,* which is often defined as the feeling of being mentally slow, fuzzy, or “spaced out”. Brain fog affects one’s ability to think or concentrate (9) and has been reported to have a substantial impact on return to work and daily life (4,10,11).

Despite the high prevalence of brain fog in Long Covid, very few evidence-based treatments address the specific cognitive impairments related to brain fog, leaving clinicians relying primarily on clinical judgement to design treatment, and causing many individuals with the condition to struggle with daily functioning (12,13). Recent reports show mixed findings, with some studies demonstrating promising outcomes following cognitive rehabilitation therapy, while others report no significant improvements with specific neurorehabilitation interventions (14–19). The scarcity of established and appropriate interventions creates the need for clinical research that can inform therapeutic treatment for individuals affected by brain fog related to Long Covid.

Some evidence-based interventions have been found to be beneficial in treating cognitive deficits resulting from conditions similar in symptoms to Long Covid brain fog, especially in people with mild traumatic brain injuries (mTBI). Given that many aspects of the cognitive profile of Long Covid brain fog resemble the cognitive deficits experienced by people with mTBI and post-concussion syndrome (i.e., deficits in attention, memory, and executive function), the interventions used for mTBI may have the potential to serve as treatment for people with Long Covid.

Attention is a foundational cognitive ability that is necessary for all other cognitive abilities (20–22). Therefore, attention deficits often affect performance in other cognitive abilities. For example, not being able to pay attention to new information will make it impossible to remember that information later. Similarly, without proper attention given to a specific task one cannot efficiently plan the next steps, problem solve or manage two tasks at the same time (i.e., executive functions). Given that Long-Covid brain fog is typically manifested with deficits in attention, (3,23–25), and in view of the fact that attention is a foundational factor in higher level cognitive skills, it could be expected that treating attention impairments may favorably affect other high level cognitive functions, resulting in less burden from brain fog (See Fig 1 for illustration). Thus we aim to develop an intervention that will be focused on improving attention. There are several evidence-based interventions to remedy attention impairments, that include: structured restorative attention training that address underlying processes, compensatory adaptation to activity requirements allowing a gradually resumed independence, and metacognitive strategy training to improve generalization of learned skills (26). Specifically, the Attention Process Training program, 3^rd^ Version (APT-3) (27) is an evidence-based clinical program for attention training, initially developed for people with acquired brain injury. It is a theoretically - driven treatment, that targets attention training based on hierarchical repetition, compensatory adaptation, and metacognitive strategy training. It addresses different types of attention including sustained, selective, alternating, and divided attention. APT-3 has been used effectively in cognitive rehabilitation with people who had a Traumatic Brain Injury (TBI) (28–34).

**Fig. 1.**
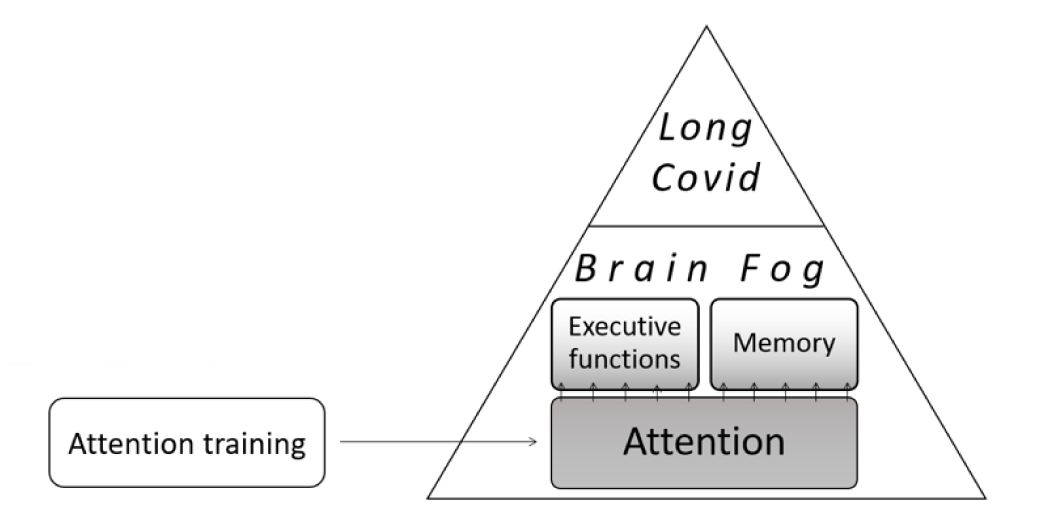
Study rationale. Targeting attention is hypothesized to result in improved executive functions and memory—core components of brain function associated with Long Covid.

The aims of this study are 1) to assess the feasibility and acceptability of using APT-3, in a sample of people with brain fog related to Long Covid, and 2) to test the efficacy of APT-3 in treating brain fog symptoms in individuals with Long Covid, compared to no treatment and to a behavioral treatment that does not target attention (music activity). It is hypothesized that a treatment targeting attention may reduce symptoms of brain fog.

## Methods

### Study Design and Timeline

This study is a randomized controlled trial with three arms: Immediate Attention Training, Music Activity group, and Delayed Attention Training. All participants have the opportunity to complete the attention training (APT-3) if desired. (See Figure 2 for a flowchart of the study).

**Fig 2.**
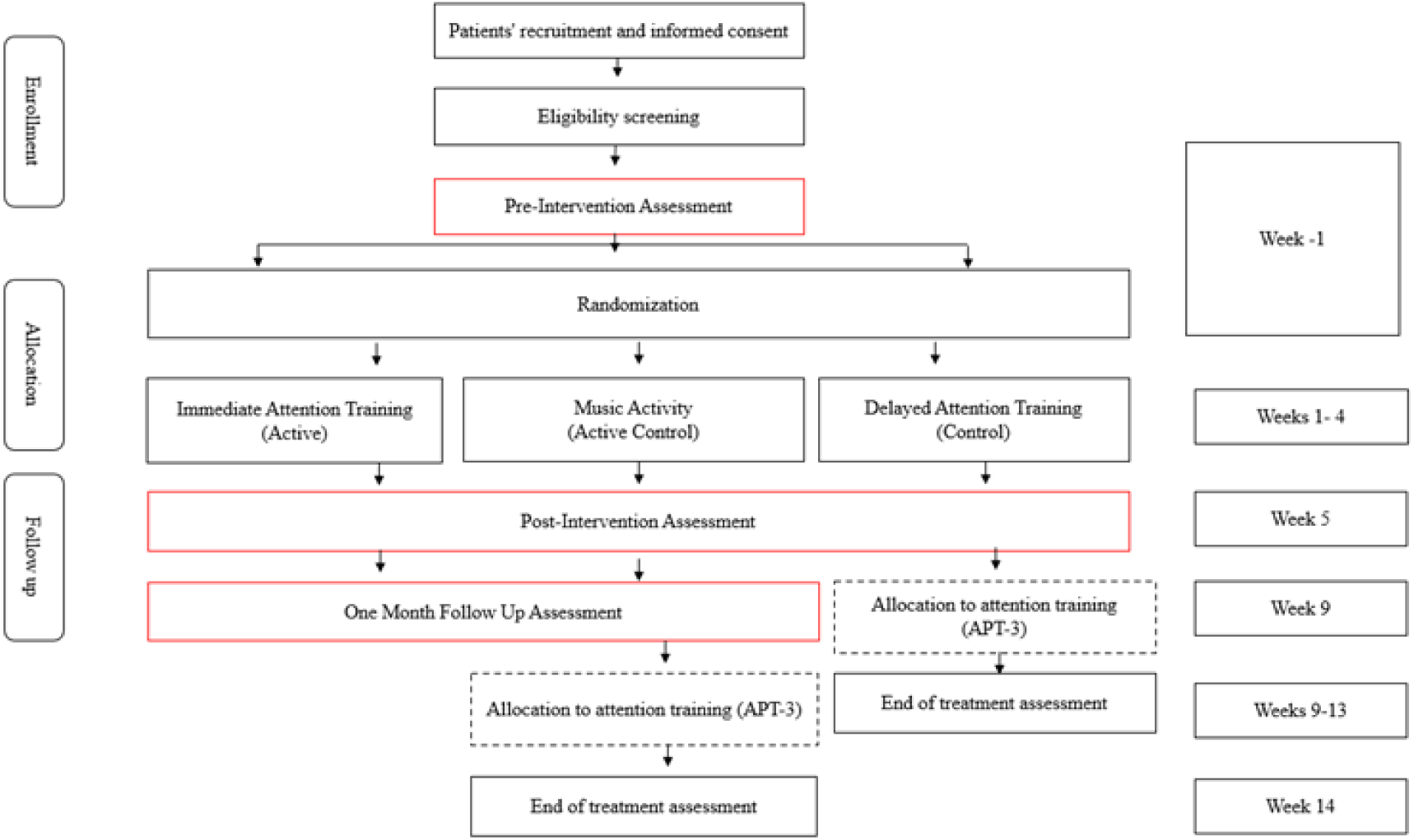
Flowchart of the study design.

All in-person study procedures were conducted at Shirley Ryan AbilityLab, Think and Speak Lab, Chicago, IL. The study has been approved by Northwestern University IRB (STU00220499). The study is registered on ClinicalTrials.gov (ID: NCT06503874).

All participants complete an initial online screening questionnaire, followed by a telephone-based eligibility interview conducted by a trained study team member. Eligible individuals who agree to participate provide informed consent prior to completion of the baseline assessment.

All participants complete an initial cognitive assessment followed by a possible 1-3 additional cognitive assessments depending on their group placement. Total study duration varies by group assignment, ranging from 5 to 14 weeks. See fig.2. Once the baseline assessment is complete, each participant is randomized to one of the following groups:

1. **Immediate Attention Training:** Participants complete a 4-week APT-3 intervention between baseline and post-intervention assessments, followed by a one-month follow-up assessment at week 9.
2. **Music Activity:** Participants engage in a 4-week music-based activity between baseline and post-intervention assessments. Following the one-month follow-up assessment at week 9. Upon completion, participants are offered 4 weeks of APT-3 and an additional post intervention assessment, extending participation up to 14 weeks.
3. **Delayed Attention Training:** Participants complete two assessments approximately four weeks apart without intervention and are subsequently offered APT-3, with post-intervention assessment at week 9 if they elect to participate.

### Participants

#### Recruitment and Eligibility

Potential participants are identified through a variety of means, including physician and therapist contacts, recruitment flyers within Shirley Ryan AbilityLab and other surrounding hospitals, support groups, websites, and ClinicalTrials.gov.

Individuals who express interest in participating in the study first complete an online screening survey on Research Electronic Data Capture System (REDCap) to assess basic eligibility criteria. A research team member then contacts potentially eligible candidates for a phone interview, in which eligibility criteria is assessed further, and information on study participation is provided.

Information collected for eligibility includes medical information (i.e., date of symptom onset, acute Covid-19 hospitalization status, perceived duration of acute Covid-19 illness (days), Covid-19 vaccination status, Long Covid symptoms and current medications). Eligible individuals who are willing to participate in the study are scheduled to review and sign the consent form and complete the baseline assessment.

The study consists of individuals, ages 18-75 years old, who are experiencing brain fog related to a diagnosis of Long Covid. Eligible participants must also meet the following inclusion criteria: 1) history of confirmed SARS-CoV-2 infection, 2) either not hospitalized for Covid care or hospitalized for immediate care following Covid for not more than 3 days, 3) subjective reports of cognitive symptoms that interfere with everyday activities, starting on or shortly after SARS-CoV-2 infection, 4) continuation or development of Long-Covid brain fog 3 months or more following the initial SARS-CoV-2 infection, with these symptoms lasting for at least 2 months and no other explanation for the symptoms, 5) ability to use a keyboard, because of the nature of some of the assessments, 6) ability to understand and communicate in English, and 7) ability to consent independently.

Participants are excluded from the study if they: 1) have been hospitalized for more than 3 days due to Covid diagnosis, 2) have pre-morbid neurological conditions that could potentially affect cognition, such as Parkinson’s Disease, Alzheimer’s Dementia, or acquired brain injury, 3) have severe depression, 4) are receiving cognitive training or physical exercise training within the prior 4 weeks, 5) are receiving chemotherapy or radiation treatment within the last 6 months, 6) have active substance abuse, 7) have any physical impairments that would affect their ability to complete the assessments (e.g., severe vision or hearing loss).

Participants are not allowed to receive any physical, occupational or speech-language therapy while enrolled in the study if related to Long Covid rehabilitation. If participants are receiving rehabilitation therapy services, they must wait 4 weeks following completion of the therapy to be enrolled in the study. This is done in order to avoid potential confounding residual effects from the therapies. If participants are taking medications or attending mental health counseling sessions, they can still be enrolled in the study, but are asked to report the treatments received and to report any dosing changes that occur during the study.

#### Sample Size

The study plans to continue participant recruitment until the 60 participants complete the study. Sample size was chosen based on best practice guidelines for conducting pilot and feasibility studies which typically suggest 12-50 participants. A formal power analysis-based sample size calculation was not performed.

#### Consenting and Retention

Eligible individuals are provided with a copy of the informed consent form via REDCap, a secure web platform used for managing research data and surveys, after completing both the online screening and phone eligibility interview. Participants are asked to review the consent form at their convenience and retain a copy for their records.

Formal consent is obtained in person at the first study visit, prior to initiating other study procedures. This process ensures that participants can ask questions and receive clarification directly from the investigator before signing the consent form and beginning the baseline assessments. Participants are provided with a copy of their signed consent form.

Each participant receives monetary compensation for their participation in the assessment sessions to offset their time and travel expenses. They are compensated $50 for the first assessment and $75 for all subsequent assessment sessions. The study timeline and expectations of time commitment are discussed prior to their consent to participate in the study. The expected dates of in-person participation are also communicated during their first assessment session. In addition, emails are sent one week prior to their scheduled sessions, as reminders to assist in maintaining adherence to the program and schedule.

#### Interventions

##### Immediate Attention Training

The APT-3 is an evidence-based, standardized computer-based training program that was designed to improve attention skills that underlie higher level cognitive processes, such as executive functions and memory (31). The APT-3 is divided into multiple sections addressing different types of attention (sustained, selective, working memory, suppression, alternating). Each week a group of 5 tasks is created by the speech-language pathologist (SLP) based on the individual reported attention challenges from the Attention Questionnaire. For example, some individuals report difficulty with maintaining attention while others only experience difficulty with focusing while distractions are present. The program of activities that are completed during the in-person session is identical to the same group of activities they complete during the week for their home sessions. Difficulty of the tasks increase progressively each week as the participants improve in their skills. APT-3 treatment is administered by a trained and certified speech-language pathologist. When the participant returns the following week, the SLP adjusts the program as needed. If the accuracy of the task is above 90%, the task is replaced with a more challenging task. If the accuracy is between 50% and 90%, the same task continues for the next week, and if accuracy is below 50%, the task is removed unless the participant indicates that they prefer to keep the more challenging task.

##### Music Activity

The Music Activity group receives enhanced exposure to music, an activity which has not been associated with improved attention skills. The Active Control group mirrors the same schedule and process as the Active Attention group; however during the 30-minute session, they listen to music and watch music videos from pre-determined playlists of their choosing (i.e. Broadway, rock, classical, etc.).

Both the APT-3 and music activity are delivered with the identical dosing and scheduling expectations. They both employ a hybrid model that combines in-person therapy sessions at Shirley Ryan AbilityLab and practice sessions which are completed independently at home. Overall, they participate in computerized training 30 minutes per day, 5 days per week, for 4 weeks (10 hours total over 4 weeks). This includes 1 day-per-week of in-person training, during which a licensed SLP reviews the participants’ tasks during the prior week, and engages in metacognitive discussions regarding their participation, self-awareness, and analysis of the tasks during the week. Questions involved in the discussion may include “What was your brain doing when you have all these errors at the beginning?”, “How hard did your brain work during those sessions?”, “Did you notice any triggers that made your brain wander during the task?”. Each group receives the same amount of time with the programs and with the therapist, with the same metacognitive discussions. The sole difference between the group interventions is the nature of the actual tasks that occur during that 30-minute time period.

#### Comparator Rationale

To perform an evaluation of efficacy of the APT-3 (attention training) for individuals experiencing brain fog resulting from Long Covid, we designed two comparator conditions: delayed attention training and a music activity. These comparator groups were selected to enable for the evaluation of a non-attention focused treatment as well as no treatment at all.

##### Delayed Attention Training Group

The use of this “waitlist control group” enables us to evaluate the natural progression of symptoms over time in the absence of any intervention. Use of this group helps to control for the effects of spontaneous recovery and regression to the mean—particularly relevant in post-viral cognitive symptoms, which may fluctuate. Participants allocated to this group are offered the APT-3 after the waitlist period, ensuring ethical equity in access to the intervention.

##### Music Activity Group

This condition involves participation in a structured music-based listening activity, matched in total duration and time of therapist contact to the APT-3 sessions. This activity was selected to control for non-specific therapeutic effects such as social interaction, cognitive engagement, and time spent on a guided task. Music-based activities have been shown to support general cognitive well-being (35,36) but are not designed to directly target attention-specific mechanisms in the way APT-3 does. Comparing APT-3 to this active condition enables us to isolate the attention-specific benefits of the intervention, beyond general engagement or placebo effects.

#### Discontinuation Criteria

Participants may discontinue their assigned intervention under the following conditions:

- **Participant Request**: Participants may withdraw from the study or stop the intervention at any time without penalty or loss of benefits.
- **Adverse Events or Discomfort**: If a participant experiences worsening symptoms that may be attributed to the intervention, the intervention may be paused, modified, or discontinued.
- **Investigator Judgment:** The intervention may be discontinued if, in the judgment of the investigators, continuing the intervention is not in the best interest of the participant (e.g., due to emerging health conditions or changes in eligibility status).

#### Outcome Measures

Primary outcomes assess feasibility and acceptability, including recruitment, retention, adherence, and participant satisfaction. Secondary outcomes evaluate preliminary efficacy using objective neuropsychological measures and validated self-report instruments administered at baseline, post-intervention, and one-month follow-up (Table 1).

**Table 1.** Objective and subjective measures at assessments and weekly sessions.

| Target construct | Measurement tool | Baseline Assessment | Post Treatment Assessment | Maintenance Assessment |
| --- | --- | --- | --- | --- |
| *Treatment* | Entry Interview | X |  |  |
|  | Exit Interview |  | X |  |
|  | Treatment Acceptability/Adherence Scale (TAAS) | X |  |  |
| *Attention-Objective Measures* |  |  |  |  |
| *1.Sustained Attention* | • Conners CPT-3<br>• Paced Auditory Serial Addition Test (PASAT)<br>• Forward digit span subtest of the WAIS-III<br>• Trials A from the Delis-Kaplan Executive Function System (D-KEFS) | X | X | X |
| *2.Attention control of executive function* | • Backward digit span subtest of the WAIS-III<br>• Trails B from the Delis-Kaplan Executive Function System<br>• Stroop test from the Delis-Kaplan Executive Function System (D-KEFS) | X | X | X |
| *3.Attention control of learning and memory* | • California Verbal Learning Test (CVLT) | X | X | X |
| *Scales* |  |  |  |  |
| *Attention-self reported* | • The Attention Questionnaire | X |  |  |
| *Depression* | • Patient Health Questionnaire-9 | X |  |  |
| *Anxiety* | • Generalized Anxiety Disorder Assessment (GAD-7) | X |  |  |
| *Fatigue* | • Fatigue Scale for Motor and Cognitive Functions (FSMC) | X |  |  |
| *Self-Efficacy* | • Self-report Efficacy scale | X | X | X |

#### Primary Outcomes

To assess the *feasibility and acceptability* of the intervention, we track recruitment and retention rates. In addition, participants complete entry and exit interviews, as well as the Treatment of Acceptability/Adherence Scale (TAAS)(37) prior to beginning the APT-3 and music activity. These measures are designed to capture qualitative and quantitative information regarding participants’ experiences and the overall feasibility and acceptability of completing the program.

#### Secondary Outcomes

To measure the *efficacy* of the APT-3 the following assessments are conducted at three points: pre-intervention (baseline), post-intervention, and one-month follow-up. Objective measures of attention include the Conners Continuous Performance Test – 3rd Edition (CPT-3)(38), Paced Auditory Serial Addition Test (PASAT) (39), Digit Span subtest from the Wechsler Adult Intelligence Scale (WAIS-III) (40) , Trail Making and Stroop subtests from the Delis-Kaplan Executive Function System (D-KEFS) (41), and California Verbal Learning Test (CVLT) (42). Self -reported measures of attention include the Attention Questionnaire (AQ) (31). In addition, participants also complete self-report questionnaires for depression through the Patient Health Questionnaire-9 (PHQ-9) (44), efficacy through a Self-Efficacy Scale, modified from the University of Washington Self-Efficacy Scale (43), anxiety through the Generalized Anxiety Disorder Assessment-7 (GAD-7) (45), and fatigue through the Fatigue Scale for Motor and Cognitive Functions (FSMC) (46)(See table 1).

#### Assignment of Interventions Randomization and Blinding

Participant randomization into the three study arms is conducted using a list generated via Research Randomizer (https://randomizer.org/). Only the principal investigator, the intervention provider, and the participant are aware of the randomization sequence and group assignments. The investigator completing the assessments is blinded. The team of investigators regularly remind and emphasize to the participants the importance of not revealing any information about their group assignment, the interventions, or any related experience to the team member completing the assessments in order to maintain blinding of the assessor.

#### Statistical Analysis

Data is collected digitally using Northwestern University’s REDCap system or, when necessary, initially recorded on paper forms and subsequently entered into REDCap. All data is securely stored on encrypted, cloud-based servers maintained by the Shirley Ryan AbilityLab. Access to identifiable data is strictly limited to authorized study team members, and all personnel are trained in data privacy and confidentiality procedures. Data quality is monitored regularly using range checks and routine audits to identify and address any discrepancies.

Analysis of study data will be conducted after completion of all maintenance (4 weeks post APT-3 completion) assessments. Descriptive statistics will be used to summarize the first aim, feasibility/acceptability outcomes, including recruitment and retention rates, self-reported acceptability of the intervention, measured by the TAAS scale, and participant feedback collected through entry and exit surveys. In addition, interviews conducted with participants before and after the intervention will be analyzed using qualitative methods to identify key themes.

For the second aim, which examines the efficacy of APT-3 in improving attention, a repeated measures ANOVA will be used to compare change scores (post–pre) across the Immediate APT-3, Delayed APT-3, and Music Activity groups. Depression, fatigue, and anxiety- known to influence attention, will be included as covariates. To assess whether treatment gains are maintained four weeks after completion, an additional repeated measures ANOVA will be conducted using post-treatment and follow-up scores. Analyses will be limited to participants who completed the intervention. Effect sizes and confidence intervals will be reported to inform the design and sample size calculations of future trials. All analyses will be conducted using the most recent versions of SPSS and JASP. Significant results will be followed by post-hoc analyses with Bonferroni corrections.

#### Ethics and Dissemination

There are no major risks expected for participants in this study. Participants may become frustrated, tired, or bored during the assessment and the interventions. Some adverse events may include changes to participants’ mental or cognitive status affecting their daily life, such as taking time off work, self-medicating, or increase in pain. The study team is trained to respond to indications of unease with support and comfort. All adverse events that are reported to the investigators are reported to the IRB. Any changes to the study protocol, after it is approved by the IRB are submitted to the IRB for approval. Changes are submitted through the online portal which we use for all IRB communication. Northwestern IRB can audit the study at any point, and the team is prepared for any such audit. The audit checklist is used in preparation and ongoing with the study to ensure all areas meet the criteria for any audit.

## Dissemination

The investigators plan to communicate the study results at the organizational, community, and national levels, through posters, presentations and peer-reviewed journal articles. These presentations may include support groups, rehabilitation conferences, and other medical forums. At the organizational level, this may include presenting to the physicians and therapists at their monthly continuing education conferences. At the community level, this may include surrounding hospitals and clinics. At the National level, it may include conferences of the American Congress of Rehabilitation Medicine, and other rehabilitation organizations. A formal report of our findings will also be published.

## Discussion

Brain fog is a prevalent and disabling symptom of Long Covid, yet evidence-based cognitive rehabilitation approaches remain limited and inconsistent. Although emerging studies suggest that cognitive rehabilitation may benefit some individuals with Long Covid, variability in findings highlights the need for well-designed trials that target specific cognitive mechanisms. This study addresses this gap by evaluating an attention-focused intervention grounded in established cognitive rehabilitation theory.

Attention was selected as the primary treatment target due to it’s foundational role in supporting higher-order cognitive functions, including memory and executive functioning. The use of Attention Process Training-3 (APT-3) is informed by its theoretical framework and demonstrates effectiveness in populations with acquired brain injury, particularly mild traumatic brain injury, which shares a similar cognitive profile. This focus allows the study to examine whether remediation of attention may contribute to broader improvements in cognitive efficiency and daily functioning.

The randomized controlled design, incorporating both an active comparator and a delayed-treatment condition, strengthens the study’s ability to distinguish attention-specific effects from non-specific therapeutic influences and natural symptom variation. In addition to preliminary efficacy outcomes, the inclusion of feasibility and acceptability measures provides information regarding the practicality of implementing structured cognitive rehabilitation in individuals with brain fog related to Long Covid, an essential step for informing future clinical trials and translational efforts.

## Conclusion

This study will provide initial evidence regarding the feasibility, acceptability, and preliminary efficacy of attention-focused cognitive rehabilitation for individuals experiencing brain fog related to Long Covid. By targeting attention, the intervention has the potential to improve cognitive processes and daily functioning, addressing a critical gap in current care for this population. Findings from this trial will inform the design of larger, adequately powered studies and contribute to the development of evidence-based strategies for managing cognitive impairments in Long Covid, ultimately supporting more effective clinical care and improved patient outcomes.

### Trial Status

Recruitment started in March 2024 and will be completed early 2026. Final testing sessions to be completed for those participants should be finished by the end of March 2026. The database is continuously updated and prepped for analysis. This protocol version date is 9/17/2024.

## Data Availability

Data will be available upon reasonable request from the corresponding author following completion of the study.

## Acknowledgements

We thank Rachel Hitch for conducting and scoring the assessments.

## Authors contributions

Conceptualization and development of the study, KM, SCZ, LRC, ER. SCZ and ER secured funding. KM managed recruitment, scheduling, and provided interventions. KM and SCZ drafted the manuscript, with LRC and ER providing edits. All authors approved the final manuscript.

## Funding

This publication is supported by the Administration for Community Living (ACL), U.S. Department of Health and Human Services (HHS) and by the Kiwanis Neuroscience Research Foundation. The contents are those of the authors and do not necessarily represent the official views of, nor an endorsement, by ACL/HHS, or the U.S. Government.

## Ethics Approval and Consent to Participate

This study has been approved by the Northwestern University IRB (STU00220499) and electronic written consent is obtained by all participants.

## Abbreviations

APT: Attention Process Training-3
CPT-3: Conners Continuous Performance Test
CVLT: California Verbal Learning Test
IRB: Institutional Review Board
mTBI: Mild Traumatic Brain Injury
PASAT: Paced Auditory Serial Addition Test
PHQ-9: Patient Health Questionnaire
REDCap: Research Electronic Data Capture
SLP: Speech-Language Pathologist
TAAS: Test of Acceptability and Accessibility Scale
TBI: Traumatic Brain Injury
TMT: Trail Making Test

## Notes

### Competing Interest Statement

The authors have declared no competing interest.

### Clinical Trial

NCT06503874

